# Quantifying the impact of physical distance measures on the transmission of COVID-19 in the UK

**DOI:** 10.1101/2020.03.31.20049023

**Authors:** Christopher I Jarvis, Kevin Van Zandvoort, Amy Gimma, Kiesha Prem, CMMID COVID-19 working group, Petra Klepac, G James Rubin, W John Edmunds

## Abstract

**Background:** To mitigate and slow the spread of COVID-19, many countries have adopted unprecedented physical distancing policies, including the UK. We evaluate whether these measures might be sufficient to control the epidemic by estimating their impact on the reproduction number (*R*_0_, the average number of secondary cases generated per case).

**Methods:** We asked a representative sample of UK adults about their contact patterns on the previous day. The questionnaire documents the age and location of contacts and as well as a measure of their intimacy (whether physical contact was made or not). In addition, we asked about adherence to different physical distancing measures. The first surveys were sent on Tuesday 24th March, one day after a “ lockdown” was implemented across the UK. We compared measured contact patterns during the “ lockdown” to patterns of social contact made during a non-epidemic period. By comparing these, we estimated the change in reproduction number as a consequence of the physical distancing measures imposed. We used a meta-analysis of published estimates to inform our estimates of the reproduction number before interventions were put in place.

**Findings:** We found a 73% reduction in the average daily number of contacts observed per participant (from 10.2 to 2.9). This would be sufficient to reduce *R*_0_ from 2.6 prior to lockdown to 0.62 (95% confidence interval [CI] 0.37 - 0.89) after the lockdown, based on all types of contact and 0.37 (95% CI = 0.22 - 0.53) for physical contacts only.

**Interpretation:** The physical distancing measures adopted by the UK public have substantially reduced contact levels and will likely lead to a substantial impact and a decline in cases in the coming weeks. However, this projected decline in incidence will not occur immediately as there are significant delays between infection, the onset of symptomatic disease and hospitalisation, as well as further delays to these events being reported. Tracking behavioural change can give a more rapid assessment of the impact of physical distancing measures than routine epidemiological surveillance.

**Research in context:** *Evidence before this study:* Many governments have adopted physical distancing measures to mitigate the impact of the COVID-19 pandemic. However, it is unclear to what extent these measures reduce the number of contacts and therefore transmission. We searched PubMed and medRxiv on March 28, 2020, with the terms “ (coronavirus OR COVID-19 OR influenza) AND ((school OR work) AND (closure OR holiday)) AND (contact OR mixing)” and identified 59 and 17 results, respectively. Only one study conducted in China during the COVID-19 pandemic reported a reduction in daily contacts outside the home during the period of “ lockdown”. We found no other published articles that empirically quantify the impact of these measures on age- and location-specific mixing patterns.

*Added value of this study:* By surveying adults’ behaviour in the UK during a period of stringent physical distancing (“ lockdown”) and comparing the results to previously collected data, we found a large reduction in daily contacts particularly outside the home, resulting in a marked reduction in the estimated reproduction number from 2.6 to 0.62 (95% bootstrapped confidence interval [CI] 0.37 - 0.89). This method allows for rapid assessment of changes in the reproduction number that is unaffected by reporting delays.

*Implications of all the available evidence:* Changes in human contact behaviour drive respiratory infection rates. Understanding these changes at different stages of the COVID-19 pandemic allows us to rapidly quantify the impact of physical distancing measures on the transmission of pathogens.

## Introduction

Over 600,000 cases and over 30,000 deaths from COVID-19 have been recorded worldwide as of 28th of March 2020 ^1^. In an attempt to mitigate the COVID-19 pandemic, many countries have adopted unprecedented physical distancing policies^2^. On the 23rd of March, with just over 6,000 confirmed cases, the UK Government implemented strict physical distancing measures instructing individuals to stay at home and avoid leaving their house except for essential work, to take one form of exercise a day, and to buy essential items such as food and medicines. This followed the closure of sporting events, schools, restaurants, bars, gyms and other leisure or hospitality-related businesses the previous week^3^ and an increase in social distancing among the population that had been taking place for several days before the announcement^4^.

Physical distancing interventions attempt to reduce contacts relevant to infectious disease spread between individuals. Multiple surveys have been instigated on the uptake of different physical distancing measures during this current pandemic, but these have not explicitly measured contacts between people ^5–7^. To make accurate predictions on the impact of these measures, quantitative data on relevant contact patterns is required ^8–11^. Only one previous survey—conducted in two Chinese cities, Wuhan and Shanghai, in February 2020—quantified the impact of these measures on individuals’ contact patterns during the COVID-19 pandemic ^12^. In this paper, we describe a survey of contact patterns and compliance with physical distance measures and present results from a sample of adults in the UK. We evaluate whether these measures might be sufficient to control the epidemic by estimating their impact on the reproduction number (the average number of secondary cases generated per case).

## Methods

### Ethics Statement

Participation in this opt-in study was voluntary, and all analyses were carried out on anonymised data. The study was approved by the ethics committee of the London School of Hygiene & Tropical Medicine Reference number 21795.

### Survey methodology

We commissioned the market research company Ipsos to conduct a survey of UK adults (referred to here as the CoMix survey). Adults (≥18 years) were recruited into the survey by sending email invitations to existing members of their online panel. Representativeness of the general UK population was ensured by setting quotas on age, gender, and geographical location. This cohort of individuals will be requested to answer the survey every two weeks for a total of 16 weeks to track changes in their self-reported behaviour. The first surveys were sent on Tuesday 24th March, one day after a lockdown was announced for the UK.

Participants were asked about their attitudes towards COVID-19 and the effect of physical distancing interventions, whether they or any of their household members experienced any recent symptoms, whether they were tested for COVID-19, whether they had had any contact with known COVID-19 cases and whether they were affected by physical distancing measures.

Participants reported (i) if any person in their household were advised to quarantine, isolate, or limit time in their workplace or educational facility in the preceding seven days due to COVID-19, and (ii) if they heeded the advice and isolated, quarantined, or stayed away from their workplace or educational facility. In the survey, we defined quarantine as limiting contacts and staying at home, with restricted allowance for movement outside the home after a potential exposure with a COVID-19 case. We defined isolation as completely separating from uninfected contacts, including household members, either in the home or in a health facility. To assess the impact of advice and policy changes regarding physical distancing, we asked participants to indicate if they had planned to participate in a set of events in the preceding week. For each event type, they reported (i) whether they proceeded with their plan, or (ii) if it was cancelled or they decided not to go, and (iii) the frequency of the event type in the previous seven days. Additional questions were asked about preventive behaviours, such as hand washing or wearing masks, and about the use of public transport in the previous seven days.

In addition, we asked participants to record all direct contacts made between 5 am the day preceding the survey and 5 am the day of the survey. A direct contact was defined as anyone who was met in person and with whom at least a few words were exchanged, or anyone with whom the participants had any sort of skin-to-skin contact.

For every recorded contact, participants documented the age and gender of the contact, relationship to the contact, the frequency with which they usually contact this person, whether contact was physical (skin-to-skin) or not, and the setting where the contact occurred (e.g. at home, work, school, or while undertaking leisure activities, etc), including whether contact occurred in- or outside an enclosed building. Questions on social contacts were consistent with those from the UK arm of the POLYMOD survey^13^, which was used as the baseline pre-pandemic comparison dataset. Details on survey methodology and a copy of the questionnaire used are provided as supplementary material.

### Statistical analysis

We grouped study participants and contacts into the following age bands 18-29, 30-39, 40-49, 50-59, 60-69, and 70+. Age, gender, and locations of participants were compared to the 2018 mid-year estimates provided by the UK Office of National Statistics (ONS) to assess the representativeness of the study sample^14^. We descriptively analysed answers related to symptoms, attitudes, exposure to physical distancing measures, and individual preventative measures. We present the number and percentage or mean and standard deviation where appropriate (Table 3).

We calculated the average number of social contacts per person per day overall, and stratified by age category, sex, household size, location of contact, type of contact, and day of the week. We then compared the mean total number of daily contacts by age group to POLYMOD stratified by contact location.

We calculated social contact matrices for the age-specific daily frequency of direct social contacts, adjusting for the age distribution in the study population and reciprocity of contacts, using the socialmixr package in R^15^.

As children (<18 years) were not included as survey participants, we imputed contacts for younger age groups (child-child and child-adult contacts) using the POLYMOD UK data. Specifically, for those child contact groups that were missing, we used a scaled version of the POLYMOD social contact matrix. Following previous methods developed by Klepac et al^16^, as the scaling factor, we took the ratio of the dominant eigenvalues of the POLYMOD and CoMix matrices, for all age groups present in both studies, stratified by setting. Furthermore, to reflect school closures during the collection of our survey we removed school-contacts from the POLYMOD data from our analysis.

The basic reproduction number, or *R*_0_, is the average number of secondary infections arising from a typical single infection in a completely susceptible population, and can be estimated as the dominant eigenvalue of the next generation matrix ^17^. The exact form of the next generation matrix is model dependent. For respiratory infections, such as SARS-CoV-2 (the pathogen causing COVID-19), this is usually a function of the age-specific number of daily contacts, the probability that a single contact leads to transmission, and the total duration of infectiousness. Therefore, *R*_0_ is proportional to the dominant eigenvalue of the contact matrix ^15^.

We assumed that contact patterns prior to physical distancing were similar to those observed in the POLYMOD data, and that the duration of infectiousness and the probability that a single contact leads to transmission did not change during the study period. Under these assumptions, the relative reduction in *R*_0_ is equivalent to the reduction in the dominant eigenvalue of the contact matrices. By multiplying the value of *R*_0_ prior to the interventions by the ratio of the dominant eigenvalues from the POLYMOD and CoMix contact matrices, we were able to calculate *R*_0_ under the physical distancing interventions. Prior to interventions we assumed *R*_0_ followed a normal distribution with mean 2.6 and standard deviation of 0.54 based on a meta-analysis of the literature presented in the supplementary material.

To assess uncertainty, we repeated the age imputation process by taking 10,000 bootstrapped samples from both POLYMOD and CoMix matrices. For every bootstrap sample, we calculated the ratio between the dominant eigenvalues for the sampled POLYMOD and CoMix matrices. This sampling provided a distribution of relative change in *R*_0_ from the contact patterns observed in POLYMOD and CoMix. Subsequently, we scaled the initial distribution of *R*_0_ with the distribution of bootstrap samples to estimate *R*_0_ under physical distancing interventions.

Recent results of the BBC Pandemic study ^16^ suggested a decrease of nearly 50% in the average number of contacts made by teenagers (13-18 years) compared with the POLYMOD data. We assessed the sensitivity of our results to a potential reduction in contacts over time by taking a conservative reduction of 50% between 5-18 year olds in the POLYMOD study, and repeating our approach to estimate the reduction in *R*_*0*_.

## Results

### Participants characteristics

We surveyed 1,356 UK participants who recorded 3,849 contacts. The average age of participants was 47.2 years (Standard Deviation (SD) = 15, Max = 86) and 45% (608/1,356) were female (see table 1). The average household size was 3.1 (SD = 1.2, Max = 10). Data were collected between Tuesday 24th and Thursday 26th of March 2020 inclusive. Participants were recruited from across the UK. The sample included participants from London (16.5%), North of England (16.0%), Midlands and East of England (26.5%), South of England (24.4%), Wales (4.4%), Scotland (9.8%), and Northern Ireland (2.6%), while 116 participants did not report their region (Table 1). Further details of participant demographics and the average number of contacts stratified by age, gender, household size and location are presented in Table 2. Compared to the mid-year ONS population estimates taken from 2018, individuals over 70 years and individuals between the ages of 20-29 year of age were undersampled.

**Table 1:** Participants characteristics in the CoMix survey, and comparison with 2018 mid-year UK population estimates provided by the Office of National Statistics The CoMix survey does not include children under the age of 18.

|  | Number of participants<br>(%)* | UK ONS mid-year Estimate |
| --- | --- | --- |
| Location (N = 1,240) |  |  |
| North of England | 198 (16.0%) | 23.2% |
| Midlands and East of England | 328 (26.5%) | 25.4% |
| London | 205 (16.5%) | 13.4% |
| South of England | 302 (24.4%) | 22.2% |
| Wales | 54 ( 4.4%) | 4.7% |
| Scotland | 121 (9.8%) | 8.2% |
| Northern Ireland | 32 (2.6%) | 2.8% |
| Missing | 116 | - |
| Age group (N = 1,356)** |  |  |
| 0-9 | 0 | - |
| 10-19 | 28 (2.1%) | - |
| 20-29 | 185 (13.6%) | 17.1% |
| 30-39 | 275 (20.3%) | 17.4% |
| 40-49 | 249 (18.4%) | 16.7% |
| 50-59 | 233 (17.2%) | 17.6% |
| 60-69 | 280 (20.7%) | 13.9% |
| 70+ | 106 (7.8%) | 17.3% |
| Missing | 0 | - |
| Gender (N = 1,356) |  |  |
| Males | 748 (55.2%) | 49.4% |
| Females | 608 (44.8%) | 50.6% |
| Missing | 0 | - |

\* Within group percentages. \*\*There are no individuals aged less than 18 in the survey participants therefore we only compare the percentages of age groups that are fully observed in the study from the ONS mid-year estimates.

**Table 2:** Number of recorded contacts per participant per day stratified by age, gender, household size and day of the week.

| Category | Value | Number of Participants | CoMix reported contacts<br>Mean (IQR) | POLYMOD reported contacts<br>Mean (IQR) |
| --- | --- | --- | --- | --- |
| Overall | Overall | 1356 | 2.9 (1, 4) | 10.8 (6, 14) |
|  | 18-29 | 213 | 3.1 (1, 4) | 12.1 (7, 16) |
|  | 30-39 | 275 | 3.1 (1, 4) | 11.3 (6, 15) |
|  | 40-49 | 249 | 3.2 (1, 4) | 12.0 (6, 17) |
|  | 50-59 | 233 | 3.1 (1, 4) | 9.5 (5, 13) |
|  | 60-69 | 280 | 2.5 (1, 3) | 9.0 (5, 12) |
|  | 70+ | 106 | 2.0 (1, 3) | 7.6 (4, 12) |
| Gender of participant | Female | 608 | 3.0 (1, 4) | 11.3 (6, 15) |
|  | Male | 748 | 2.9 (1, 4) | 10.2 (5, 13) |
| Household size | 1 | 431 | 2.4 (1, 3) | 7.4 (3, 11) |
|  | 2 | 363 | 2.8 (2, 3) | 10.1 (5, 13) |
|  | 3 | 207 | 4.0 (3, 4) | 11.2 (6, 15) |
|  | 4 | 96 | 4.4 (4, 5) | 12.1 (7, 16) |
|  | 6+ | 56 | 5.3 (4, 6) | 14.2 (9, 17) |
|  | Unknown | 203 | 1.8 (1, 2) |  |
| Date |  |  |  |  |
| 24th March 2020 | Tuesday | 178 | 3.1 (1, 43) | - |
| 25th March | Wednesday | 1014 | 2.9 (1, 4) | - |
| 26th March | Thursday | 162 | 2.9 (1, 3) | - |
| 27th March | Friday | 2 | 5.0 (5, 5) | - |

Thirteen participants reported having been tested for COVID-19 with seven testing positive, and two participants still waiting for their results. Forty-one participants stated they had been in contact with a known COVID-19 case. In terms of perceived risk, 26.4% (359/1356) thought that it was likely that they would develop coronavirus and 48.0% (652/1356) agreed or strongly agreed that COVID-19 would be a serious disease for them if they acquired the infection.

### Impact of physical distancing measures

Participants reported data on a total of 3,824 household members, including themselves, of whom 508 (13.2%) had been asked to quarantine and 826 (21.6%) had been asked to isolate. Nearly a quarter (921; 24.1%) of household members lived in a house with someone who had at least one symptom of fever, aches, shortness of breath, or cough. Roughly 50% of the 2,122 employed individuals had either been asked to limit their time at work, had their work closed, and/or did not visit their work in the preceding 7 days (Table 3). Of those household members who attend educational establishments 67.2% (818/1217) had their institution closed with 63.3% not visiting during the previous 7 days.

**Table 3:** Indicators of adherence with public health interventions and behaviour changes reported by participants. Table 3 shows compliances with different social distancing measures due to COVID-19. N symptoms shows the total number of household members who were living in a household where someone had any of the following symptoms: (fever, aches, shortness of breath, cough), and how many individuals reported having COVID-19 symptoms themselves. The column *Asked to* refers to the total number of people who reported being asked to quarantine or isolate. The column *Have been in* shows the total number of people who reported having been in quarantine or isolation for at least one day in the seven days before the survey.

| Measure | Asked to | Have been in | At least | with COVID-19 symptom |
| --- | --- | --- | --- | --- |
| Quarantine (N = 3824) | 508 (13.2%) | 778 (20.3%) | Living in a household | 921 (24.1%) |
| Isolation (N = 3824) | 826 (21.6%) | 1,264 (33.1%) | People | 462 (12.1%) |

| Setting | Asked to limit time | Reported as closed | Did not visit |
| --- | --- | --- | --- |
| Work<br>N = 2122 | 1,006 (47.4%) | 996 (46.9%) | 1149 (54.1%) |
| School or University<br>N = 1217 | 651 (47.4%) | 818 (67.2%) | 771 (63.3%) |

| Event | Intended to visit | Visited | Cancelled | Chose not to visit |
| --- | --- | --- | --- | --- |
| Concert | 111 | 6 (5.4%) | 57 (51.3%) | 20 (18.1%) |
| Cinema | 133 | 11 (8.3%) | 54 (40.6%) | 43 (32.3%) |
| Sporting event |  |  |  |  |
| Participant | 105 | 14 (13.3%) | 46 (43.8%) | 33 (31.4%) |
| Attendee | 100 | 9 (13.3%) | 54 (54.0%) | 20 (20.0%) |
| Restaurant | 271 | 28 (28.7%) | 118 (43.5%) | 100 (36.9%) |
| Religious event | 105 | 14 (13.3%) | 46 (43.8%) | 33 (31.4%) |
| Pub | 366 | 105 (28.6%) | 119 (32.5%) | 24 (6.6%) |
| Supermarket | 1127 | 967 (85.8%) | 28 (2.5%) | 112 (10.0%) |

There were clear suggestions that physical distancing in the previous week had impacted planned activities for survey participants with 32.5% of participants having to cancel plans to visit a pub (Table 3). Contrastingly, only a small percentage of participants (2.5%) who intended to go to the supermarket were unable due to COVID-19.

### Contact patterns

The mean number of physical and non-physical contacts per person measured during this study was 2.9 (IQR = 1-4) which was 73.1% lower than was measured in POLYMOD (10.8; 6-14). The reduction in mean contacts between POLYMOD and CoMix was consistent across age, gender, and household size (Table 2). The respective social contact matrices (including physical and non-physical contacts) also reflected a much lower number of mean contacts across the age strata as presented in Figure 1.

**Figure 1:**
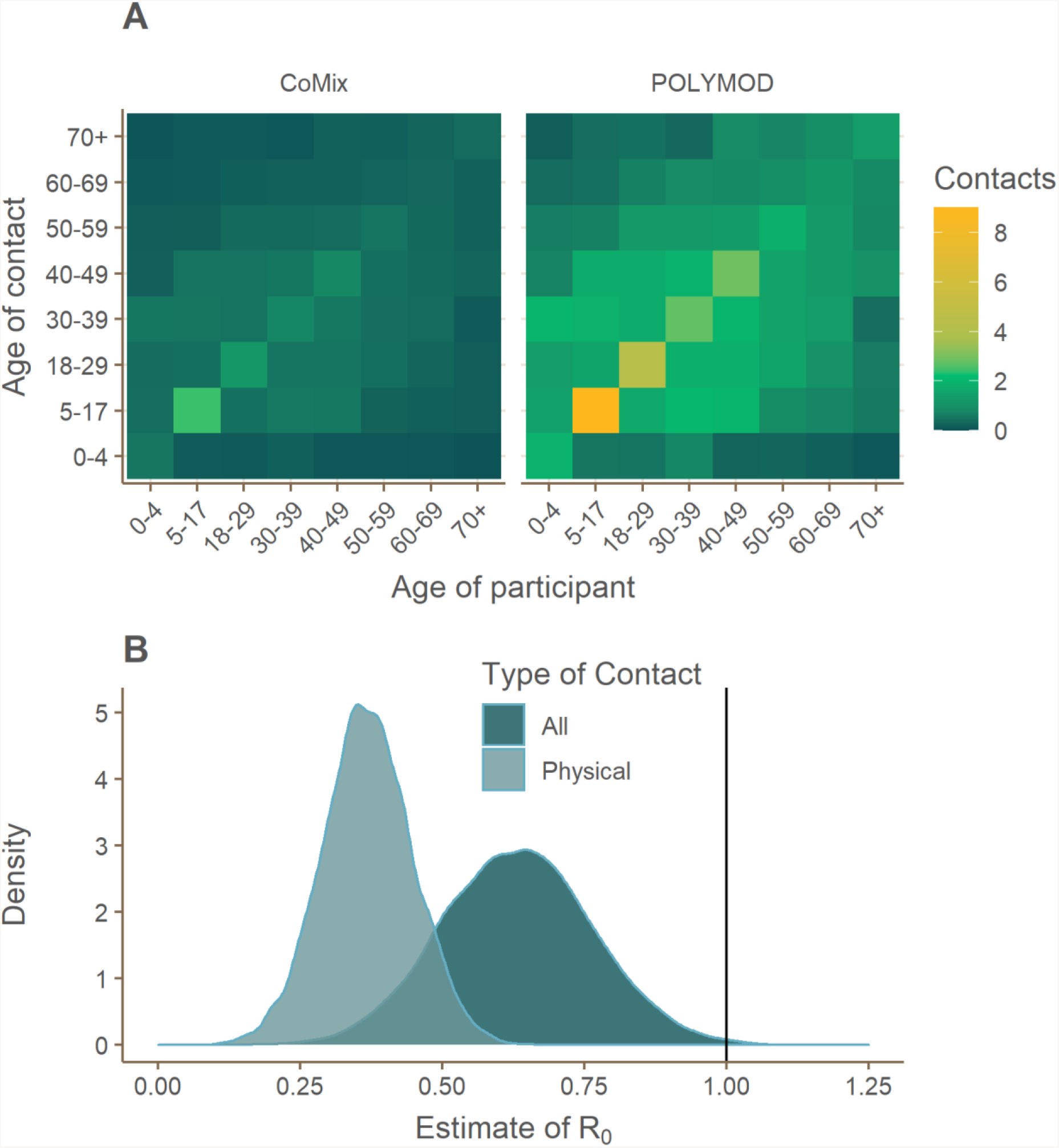
Comparison of CoMix and POLYMOD contacts matrices and estimated reduction in reproduction number due to physical distancing for all and physical contacts separately. A: Social contact matrices showing the average total number of daily reported contacts made by participants in different age groups with individuals in other age groups, with results shown for all contacts reported in the CoMix and POLYMOD data. Participants’ contacts in CoMix for age groups 0-4 and 5-17 are imputed using the POLYMOD data. B: The estimated value of R_0_ at the time of the survey, assuming values of R_0_ ∼ Norm(2.6, sd = 0.54) prior to physical distancing reducing all contacts.

The majority of contacts (57.6%) occurred at home, contrasting with 33.7% reported in the POLYMOD survey. Figure 2 displays the average number of contacts across age groups for all, physical, home, work, school, and other contacts. The matrices are consistent with the majority of contacts being in the home, with work, and other contributing very little to the overall number of contacts.

**Figure 2:**
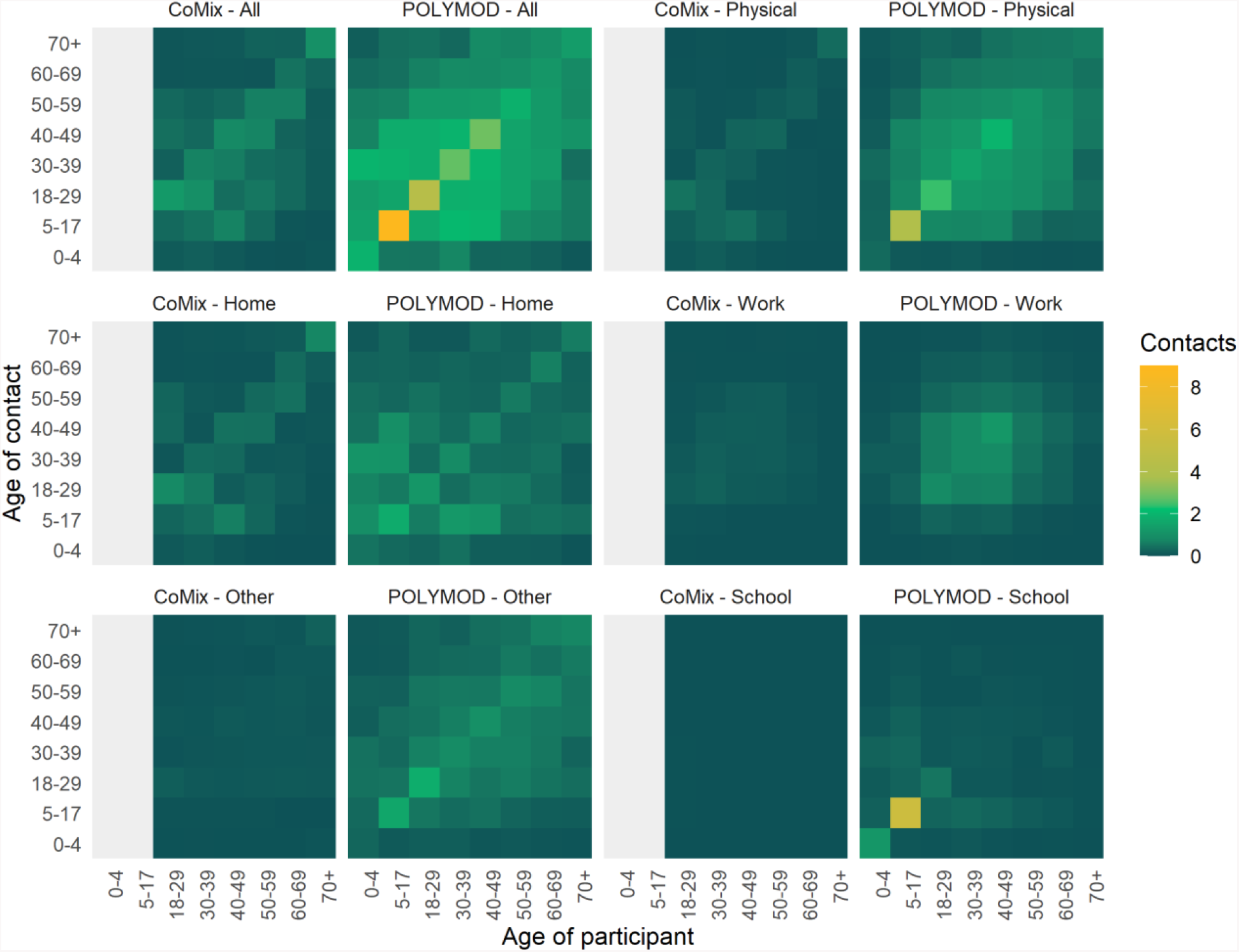
Contact matrices for all reported contacts made in different settings, comparing CoMix to Polymod.

### Estimated the basic reproduction number of COVID-19 under physical distancing

We estimated the current *R*_0_ under physical distancing measures to be 0.62 (95% confidence interval [CI] 0.37 - 0.89) based on all types of contact (Figure 1). Based on physical contacts only, we estimated R_0_ to be 0.37 (95% CI = 0.21 - 0.52). The average pre- to post-intervention ratio in R_0_ was 0.24 (min =0.21, max = 0.27) for all contacts and 0.14 (min = 0.12, max = 0.17) for physical contacts only. Based on these values, the physical distancing measures would have reduced the mean estimate of *R*_0_ to below one even if the initial *R*_0_ had been as high as 3.6 assuming all contacts are equally risky, or 4.2 assuming only physical contacts result in transmission.

In a sensitivity analysis, reducing contacts made by 5-17 year olds by 50% resulted made little difference to the results. Under this assumption the estimated value of *R*_0_ for all contacts would be 0.69 (95% CI 0.42 to 0.98) and 0.37(CI 0.22 - 0.53) if physical contacts alone result in transmission.

## Discussion

The measures introduced by the UK Government appear to have high levels of uptake among participants and have resulted in very large (73%) reductions in the total number of contacts. If similar changes are observed across the UK population, we would expect the basic reproduction number to now be below 1 (0.62; 95% CI 0.37 - 0.89), and that these physical distancing measures will lead to a decline in cases in the coming weeks. However, this projected decline in incidence will not result in an immediate decline in reported cases, as there are significant delays between infection and the onset of symptomatic disease and hospitalisation, as well as further delays to these events being reported. Hence, routine surveillance data are unlikely to show a decline in cases for some time. However, by directly measuring individuals’ contact patterns and estimating the corresponding basic reproduction number, we are able to rapidly quantify the impact of physical distancing on transmission.

The total number of daily contacts (mean of 3.1 per person) was significantly reduced compared to patterns previously estimated in the POLYMOD study (10.7; excluding children <18 years old) and more recently by the BBC Pandemic study (10.5; excluding under 13-year-olds)^16^. The observed reduction appears to be unlikely due to chance given the large difference in average contacts, and is consistent with a recent study conducted in Wuhan, China that estimated a reduction in the average number contacts per day from 14.6 prior to the outbreak to 2.0 under physical distancing interventions ^12^. While we are unaware of any directly comparable data from the UK, our findings are certainly consistent with other reports from the UK of a dramatic reduction in social contacts, with, for example, only half of respondents in one survey reporting having the house at all in the past 24hrs^4^.

There are several limitations to this survey. Asking individuals to report their contacts from the day before may result in recall bias. Moreover, individuals who are adhering to physical distancing measures may have been more likely to respond to this survey, potentially resulting in selection bias and in an overestimate of the impact of these measures. We were not able to sample any children, so child-child contacts had to be imputed from comparison with a previous survey.

We were not able to quantify any additional effect from the interventions on transmission, such as reduction in infectiousness by increased handwashing. In addition, we were not able to calculate the net reproductive number, *R*_0_, as we did not account for the proportion of the population that is no longer susceptible. These could all reduce the net reproductive number to values lower than estimated in our analysis.

Our analysis assumed that direct contacts are an appropriate proxy for effective contacts, and thus that transmissibility is equal across age-groups (e.g. contact between a single infected child and susceptible adult is as likely to result in transmission as contact between a single infected adult and a susceptible adult). We further assume that the reduction in non-school contacts in children is similar to that observed in adults. Furthermore, we assume that the contact patterns prior to interventions are consistent and of similar magnitude. A recent study has found significantly lower numbers of contacts reported by teenagers compared with the POLYMOD survey^16^. Decreasing mixing among 5-17 years by 50%, whilst reducing the magnitude of reduction in R_0_, did not affect the qualitative conclusions from the analysis.

This study is planned to continue in the UK for the next 15 weeks. Future analyses will be able to explore changes in contact patterns during different interventions and may provide early warning signs of changes in contact patterns due to interventions being lifted or decreasing adherence with restrictions.

## Conclusions

We have shown that behavioural monitoring can give a rapid insight into transmission of COVID-19 and have provided the first evidence that the restrictions adopted by the UK government have led to a decrease in transmission of COVID-19.

## Data Availability

The data for the contact matrices will be made available in the coming days.

## Acknowledgements

This project was funded by the European Union’s Horizon 2020 Research and Innovations Programme - project EpiPose (Epidemic Intelligence to Minimize COVID-19’s Public Health, Societal and Economical Impact, No 101003688).

WJE, GJR, CIJ, and KvZ, conceived of and designed the study. CIJ, KvZ, and WJE conceived of the analysis. CIJ, KvZ, AG, and KP conducted the analysis. CIJ and KvZ wrote the manuscript with input and guidance from WJE, AG, KP, GJR, and PK. The CMMID COVID-19 working group members contributed to processing, cleaning, and interpretation of data, interpreted the study findings, contributed to the manuscript, and approved the work for publication. All authors interpreted the findings, contributed to writing the manuscript, and approved the final version for publication.

CIJ and AG were funded by the Global Challenges Research Fund (ES/P010873/1). KvZ (Elrha’s Research for Health in Humanitarian Crises [R2HC] Programme, UK Government [Department for International Development], KP was funded by the National Institute for Health Research (NIHR; 16/137/109). PK was funded by the Bill & Melinda Gates Foundation (INV-003174). GJR was funded by the National Institute for Health Research Health Protection Research Unit (NIHR HPRU) in Emergency Preparedness and Response at King’s College London in partnership with Public Health England (PHE), in collaboration with the University of East Anglia and Newcastle University. The views expressed are those of the author(s) and not necessarily those of the NHS, the NIHR, the Department of Health and Social Care or Public Health England

We would like to acknowledge the other members of the London School of Hygiene & Tropical Medicine CMMID COVID-19 modelling group, who contributed to this work. Their funding sources are as follows: Megan Auzenbergs and Kathleen O’Reilly (Bill and Melinda Gates Foundation, OPP1191821); Graham Medley (NTD Modelling Consortium by the Bill and Melinda Gates Foundation (OPP1184344); Jon C Emery and Rein M G J Houben (European Research Council Starting Grant, Action Number #757699); Nicholas Davies (NIHR HPRU-2012-10096); Emily S Nightingale (Bill and Melinda Gates Foundation, OPP1183986); Wellcome Trust, and NIHR); Stefan Flasche (Sir Henry Dale Fellowship 208812/Z/17/Z); Thibaut Jombart (Research Public Health Rapid Support Team, NIHR Health Protection Research Unit Modelling Methodology); Joel Hellewell, Sam Abbott, James D Munday, Nikos I Bosse and Sebastian Funk (Wellcome Trust 210758/Z/18/Z); Fiona Sun (NIHR; 16/137/109); Akira Endo (The Nakajima Foundation; The Alan Turing Institute); Alicia Rosello (NIHR: PR-OD-1017-20002); Simon R Procter (Bill and Melinda Gates Foundation, OPP1180644); Fiona Sun (NIHR; 16/137/109); Adam J Kucharski and Timothy W Russell (Wellcome Trust, 206250/Z/17/Z); Gwen Knight (UK Medical Research Council, MR/P014658/1); Hamish Gibbs (Department of Health and Social Care ITCRZ 03010); Quentin Leclerc (Medical Research Council London Intercollegiate Doctoral Training Program studentship (grant no. MR/N013638/1); Billy J. Quilty, Charlie Diamond, Yang Liu and Mark Jit National Institute for Health Research NIHR; 16/137/109). Yang Liu and Mark Jit (Bill & Melinda Gates Foundation INV-003174); Samuel Clifford (Sir Henry Dale Fellowship 208812/Z/17/Z); Carl A.B. Pearson (NTD Modelling Consortium by the Bill and Melinda Gates Foundation OPP1184344); Rosalind M. Eggo (Health Data Research UK MR/S003975/1); Arminder K Deol; This research was partly funded by the NIHR (16/137/109) using aid from the UK Government to support global health research. The views expressed in this publication are those of the author(s) and not necessarily those of the NIHR or the UK Department of Health and Social Care. We would also like to thank Paula Bianca Blomquist for thoughtful critique and discussions during this analysis.

